# COVID-19 Prediction using Genomic Footprint of SARS-CoV-2 in Air, Surface Swab and Wastewater Samples

**DOI:** 10.1101/2022.03.14.22272314

**Authors:** Helena M. Solo-Gabriele, Shelja Kumar, Samantha Abelson, Johnathon Penso, Julio Contreras, Kristina M. Babler, Mark E. Sharkey, Alejandro M. A. Mantero, Walter E. Lamar, John J. Tallon, Erin Kobetz, Natasha Schaefer Solle, Bhavarth S. Shukla, Richard J. Kenney, Christopher E. Mason, Stephan C. Schürer, Dusica Vidovic, Sion L. Williams, George S. Grills, Dushyantha T. Jayaweera, Mehdi Mirsaeidi, Naresh Kumar

## Abstract

**Importance:** Genomic footprints of pathogens shed by infected individuals can be traced in environmental samples. Analysis of these samples can be employed for noninvasive surveillance of infectious diseases.

**Objective:** To evaluate the efficacy of environmental surveillance of severe acute respiratory syndrome coronavirus 2 (SARS-CoV-2) for predicting COVID-19 cases in a college dormitory.

**Design:** Using a prospective experimental design, air, surface swabs, and wastewater samples were collected from a college dormitory from March to May 2021. Students were randomly screened for COVID-19 during the study period. SARS-CoV-2 in environmental samples was concentrated with electronegative filtration and quantified using Volcano 2^nd^ Generation-qPCR. Descriptive analyses were conducted to examine the associations between time-lagged SARS-CoV-2 in environmental samples and clinically diagnosed COVID-19 cases.

**Setting:** This study was conducted in a residential dormitory at the University of Miami, Coral Gables campus, FL, USA. The dormitory housed about 500 students.

**Participants:** Students from the dormitory were randomly screened, for COVID-19 for 2-3 days / week while entering or exiting the dormitory.

**Main Outcome:** Clinically diagnosed COVID-19 cases were of our main interest. We hypothesized that SARS-CoV-2 detection in environmental samples was an indicator of the presence of local COVID-19 cases in the dormitory, and SARS-CoV-2 can be detected in the environmental samples several days prior to the clinical diagnosis of COVID-19 cases.

**Results:** SARS-CoV-2 genomic footprints were detected in air, surface swab and wastewater samples on 52 (63.4%), 40 (50.0%) and 57 (68.6%) days, respectively, during the study period. On 19 (24%) of 78 days SARS-CoV-2 was detected in all three sample types. Clinically diagnosed COVID-19 cases were reported on 11 days during the study period and SARS-CoV-2 was also detected two days before the case diagnosis on all 11 (100%), 9 (81.8%) and 8 (72.7%) days in air, surface swab and wastewater samples, respectively.

**Conclusion:** Proactive environmental surveillance of SARS-CoV-2 or other pathogens in a community/public setting has potential to guide targeted measures to contain and/or mitigate infectious disease outbreaks.

**Key Points:** *Question:* How effective is environmental surveillance of SARS-CoV-2 in public places for early detection of COVID-19 cases in a community?

*Findings:* All clinically confirmed COVID-19 cases were predicted with the aid of 2 day lagged SARS-CoV-2 in environmental samples in a college dormitory. However, the prediction efficiency varied by sample type: best prediction by air samples, followed by wastewater and surface swab samples. SARS-CoV-2 was also detected in these samples even on days without any reported cases of COVID-19, suggesting underreporting of COVID-19 cases.

*Meaning:* SARS-CoV-2 can be detected in environmental samples several days prior to clinical reporting of COVID-19 cases. Thus, proactive environmental surveillance of microbiome in public places can serve as a mean for early detection of location-time specific outbreaks of infectious diseases. It can also be used for underreporting of infectious diseases.

## INTRODUCTION

The COVID-19 pandemic caused by the severe acute respiratory syndrome coronavirus 2 (SARS-CoV-2) has resulted in 485 million cases and 6.14 million deaths worldwide as of April 1, 2022 [1]. It has impacted societies, economies, cultures, ecologies, and politics globally in an unprecedented manner, and the control and management of the pandemic has been challenging. Limited human testing, especially among asymptomatic cases, and mutation of the virus further exacerbate this challenge. As infected individuals travel across space, they shed the virus through bodily fluids and exhalation (in the form of air droplets). As a result, the virus makes its genomic footprint in the environment, including in air, on surfaces and in wastewater. The virus can survive in these environments for hours to weeks, especially indoors, depending on the surface type and meteorological conditions [2, 3]. Thus, physical contact and/or inhalation of aerosols contaminated with SARS-CoV-2 are considered as main routes of contracting the virus [3, 4]. Quantifying the genomic footprints of the virus in the environmental samples offers a unique opportunity for proactive disease surveillance, which can guide targeted management strategies.

Environmental monitoring of pathogens is non-invasive, cost-effective and can be conducted in public spaces. Thus, it can provide early indications of community level infection and early warnings of disease outbreaks more effectively than human surveillance methods (i.e. testing and tracing infected individuals). Given the multimodal transmission of SARS-CoV-2, researchers have been monitoring the virus in wastewater [5-7], on surfaces [8] and in the air [9]. Each medium has its unique advantages and disadvantages. For example, wastewater monitoring can localize the source of infection to a building, a neighborhood, and/or a community. Research shows that the concentrations of SARS-CoV-2 found in wastewater samples correspond with COVID-19 case incidence rate [10-14]. Although there is some consistency in the association between SARS-CoV-2 in the wastewater samples and COVID-19 cases, this association can vary with respect to scale (ranging from building to community sewage plant) and time-lag between COVID-19 case reporting and SARS-CoV-2 detected in the wastewater sample [13]. For example, a recent study shows that the four-day lagged SARS-CoV-2 concentration in the wastewater samples had the strongest association with COVID-19 cases at a university campus in 2020-21 and this association varied across different parts of the campus [13]. While wastewater surveillance provides an early indication of community level infection, it is challenging to trace potential location(s) of cases in the absence of building specific monitoring of wastewater. Moreover, the virus concentration in the wastewater can be influenced by dilution, distance and time between the source and sample site, time of sampling, and area of the wastewatershed.

Although SARS-CoV-2 in air, surface and wastewater samples has been detected [4, 15, 16], limited data are available on their relative comparison and efficacy in COVID-19 case prediction rate (see Table S1). This paper aims to address this research gap by using the data from a controlled experimental design. We compare time- and building-matched concentrations of SARS-CoV-2 in daily air, surface and wastewater samples in a student dormitory (of about 500 students) for three months in Spring 2021 at the University of Miami, Coral Gables, FL, USA campus (UM). Regular screening of the student population for COVID-19 by building allowed for a direct comparison of environmental detection of SARS-CoV-2 coming from the same source of infected individuals.

## MATERIALS AND METHODS

### Experimental strategy

We used a prospective control design for this research. We selected a campus site (Y-leg of Lakeside Village, YLV) with corresponding wastewater and high traffic access points. YLV houses about 500 students and is serviced by two interconnected lobbies, C and D. This dorm is secure and can be accessed by students by swiping their university issued identification card. Based on the swipe registry data, more than 5,000 individuals (mostly students) entered YLV in a seven-day period. The wastewater from the YLV drains to a designated manhole (K). Since there are two access points to YLV, we conducted daily (active) air sampling and surface swab sampling in both lobbies. Thus, 24h air samples and swab samples of high touch surfaces, namely elevator buttons, door handles and bars, (Figure S1) corresponded to daily wastewater samples from manhole K. Random screening of students residing in the dormitory also occurred 2-3 times/week during the study period: March 2, 2021 to May 28, 2021. Methods of sample collection and analyses of these samples are detailed below. Daily air, surface swab and wastewater samples were collected between 8:00 AM and 9:00 AM Monday through Friday and 10:00 AM to 11:00 AM during weekends.

### Wastewater sampling at K-Manhole

Both grab and 24h time-paced composite samples (ISCO 6712) were collected with some overlap. The sampling team adhered to the University’s Health and Safety Office protocol for collecting, preserving, transporting and processing of these samples. Each day 250 ml of wastewater sample was collected in a prelabelled disposable bottle, and temperature, pH, salinity, dissolve oxygen, and turbidity of the sample were measured using a pre-calibrated sonde (Xylem YSI Pro DSS). The samples were transported to our laboratory in an ice cooler for processing in a BSL-2 cabinet (see SOM for details).

### Air sampling in lobby C and D of YLV

Active air sampling was conducted with impactors connected with a vacuum pump. Air sampling setup was designed strategically to capture air from all floors. We positioned air impactors between two elevator doors in both lobbies, which can potentially represent air from all floors and aerosols exhaled by all subjects entering and/or exiting the building (Fig. S2). This location had several advantages. First, everyone entering or exiting the dorm had to wait inside and outside the elevator until the elevator door opens. Thus, individuals can exhale several liters of air depending on the wait time, which will be drawn by the vacuum inlet positioned near the elevator door. Second, while in the elevator, an individual would have been inhaling and exhaling aerosols. As soon as the elevator door opens the vacuum inlet will draw air dispensed from inside the elevator due to positive pressure inside the elevator.

We used sterile 47mm polycarbonate membrane filters with a pore size of 0.4 µm in our customized impactor, connected with a vacuum pump. Filters were placed onto a new 47 cellulose absorbent pads to avoid any leakage and membrane breakage. Air was sampled for 24h at a flow rate of 10-12 L/minute. Initial and end flow rates were noted along with meteorological conditions and the concentration of airborne particulate matter of different sizes in the lobby. Samples were preserved in a sterile and pre-labelled DNA/RNase free sterile 5mL conical tube and transported to the laboratory in an ice cooler. After sample collection, impactors were cleaned with 70% isopropyl alcohol, and a new set of filters with new supporting absorbent pads were deployed for the next 24h.

### Surface sampling in lobby C and D of YLV

Daily surface swab samples were collected using a sterile a Polyester Swab with Polystyrene Handle after retrieving the air filters (deployed 24h ago) and deploying new air filters. The high touch surfaces, including elevator buttons and door handles, were swabbed using a unidirectional scrapping motion. After swabbing the surface, it was wiped and cleaned with a wet alcohol wipe. The swab tip was preserved in a pre-labelled 1.5ml DNA/RNAse free sterile conical tube. Date, time, temperature, and humidity were noted and samples were transported in an ice cooler to the laboratory. There are two entrance doors to lobby C and three entrance doors to lobby D. Thus, the surface area of the door bars, handles and elevator buttons were 578.9 square inches in lobby C and 1,136.7 square inches in lobby D.

### Processing and RNA extraction

Air and surface samples were preserved in 1 mL of DNA/RNA shield (Zymo Research). Heat inactivated human coronavirus HCoV-OC43 (ZeptoMetrix) with the known concentration between 10^5^ and 10^6^ gc/L was added to air, surface swab and wastewater samples as an RNA recovery control. All samples were processed within 3h after the collection. Wastewater sample processing included continuous mixing after adding MgCl_2_ to a concentration of 50 mM and acidification to a pH value between 4.5 to 3.5. Acidified samples were filtered through an electronegative membrane (0.45um pore size, 47mm diameter, SM: Pall HA/MCE membrane, UM: Millipore HAWP4700) [17-19]. Wastewater concentrates consisted of membranes preserved 1 mL DNA/RNA shield. After mixing in the corresponding DNA/RNA shield, air, surface swab and wastewater concentrates, 250µL aliquots were extracted using a Zymo Quick-RNA Viral Kit. The extracted RNA was eluted in 15µL of RNase free water and preserved in -80° C freezer until the V2G-qPCR analysis.

### SARS-CoV-2 quantitation

We used Volcano Second Generation qPCR (V2G-qPCR) as described in Sharkey et al.[13]. An advantage of the V2G-qPCR over RT-qPCR is that it can read both RNA and DNA templates and eliminates the cDNA synthesis step [20]. Moreover, it is less sensitive to inhibitors found in environmental samples. Three microliter aliquots of purified RNA were used in singleplex reactions to quantify the SARS-CoV-2 N3 nucleocapsid gene [21] and OC43 as a recovery control. We used Twist Synthetic SARS-CoV-2 RNA standards to quantitate SARS-CoV-2 GC in the sample. Negative controls were used to control for contamination. SARS-CoV-2 and OC43 concentrations in genomic copies (GC) per air volume, surface area wiped, or volume of wastewater were then calculated using corresponding dilution and/or concentration factors used at each step of processing.

### COVID-19 surveillance

Students residing at the YLV dormitory were randomly screened 2-3 days/week using nasal swabs which were analyzed using RT-PCR. These anonymous data on the total number of tests (performed) and COVID-19 cases by date were acquired from the UM administration.

### Statistical analysis

We conducted descriptive analyses of aggregated and disaggregated data. Data of air and surface swab samples from both lobbies were aggregated to compare them with the data from the wastewater samples. χ^2^ tests were performed to assess statistical differences across groups and a p-value of 0.05 or below was considered as significant.

## RESULTS

A total of 445 air, surface swab and wastewater samples were collected from March 2 to May 25, 2021. Of these, 165 air samples and 166 surface swab samples were collected from two lobbies of Lakeside Village (YLV) at the UM campus. A total of 114 daily wastewater samples were collected from manhole K. On 24 days, we collected both grab and 24h composite samples. On 4 days when SARS-CoV-2 was detected in composite samples it was below detection limit (LOD) in the grab samples. On 3 days it was detected in the grab samples but below LOD the composite samples. Although the mean concentration of SARS-CoV-2 in samples with above LOD (n=27) was slightly higher in the composite samples as compared to grab (917.9 genomic copies (gc)/L versus 597 gc/L on average), these differences were statistically insignificant. The concentration and frequency of SARS-CoV-2 detection in the air and surface samples collected from each of the two lobbies of YLV did not vary significantly. We detected SARS-CoV-2 in 50 (30%) of the 165 air samples. The average SARS-CoV-2 concentration in the air sample was 14.8 gc/m^3^. Of the 166 surface swab samples, SARS-CoV-2 was detected in 33 (19.9%) of them. The average concentration of SARS-CoV-2 in these samples was 16.5 gc/m^2^ surface area. Of the 114 grab and composite wastewater samples SARS-CoV-2 was detected in 57 (or 50%) of the samples and the average concentration in these samples was 1390 gc/L (Table 1).

**Table 1:** SARS-CoV-2 detection and concentration in air, surface, and wastewater samples.

| Environmental Matrix | Number of samples collected | Number of SARS-CoV-2 positive samples (% total) | $\log_e$ (SARS-CoV-2 gc) (95% Confidence Interval (CI); n) |
| --- | --- | --- | --- |
| Air (gc/m<sup>3</sup>) | 165 | 50 (30.3) | 2.7<br>(2.4 – 3.0; 50) |
| Surface (gc/m<sup>2</sup>) | 166 | 33 (19.9) | 2.8<br>(2.4 – 3.2; 33) |
| Wastewater (gc/L) | 114 | 57 (50.0) | 6.5<br>(6.2 – 6.8; 57) |
| Total | 445 | 140 (31.5) | 4.3<br>(3.9 – 4.6; 140) |

Air and surface swab samples were aggregated by day to compare against the wastewater samples. Some consistency was observed in the concentration and detection of SARS-CoV-2 across air, surface and wastewater samples. On 36 (or 46%) of 78 matched days there was complete agreement in the SARS-CoV-2 detection in all air, surface, and wastewater samples. However, on many of the days, SARS-CoV-2 was detected in the wastewater samples but not in air and surface swab samples, and vice-versa (Figure 1). For example, SARS-CoV-2 was not detected in wastewater samples on 24 days, but 17 and 14 of these 24 days the virus was detected in the air and surface swab samples, respectively (Table S2). The difference in the frequency distribution of the virus detection across different sample types was marginally significant (χ^2^ = 3.06; p ∼ 0.058). When SARS-CoV-2 was detected in 1-day lagged moving average of wastewater samples, it was also detected on 33 and 26 days in air and surface samples, but not on 21 and 28 days, respectively. The frequency distribution of the virus detection across air and surface samples when it was also detected in wastewater samples did not show statistically significant differences (χ^2^ = 3.02; p ∼ 0.082) (Table S2). This suggests a stronger agreement in the detection of SARS-CoV-2 across three different types of environmental samples.

**Figure 1:**
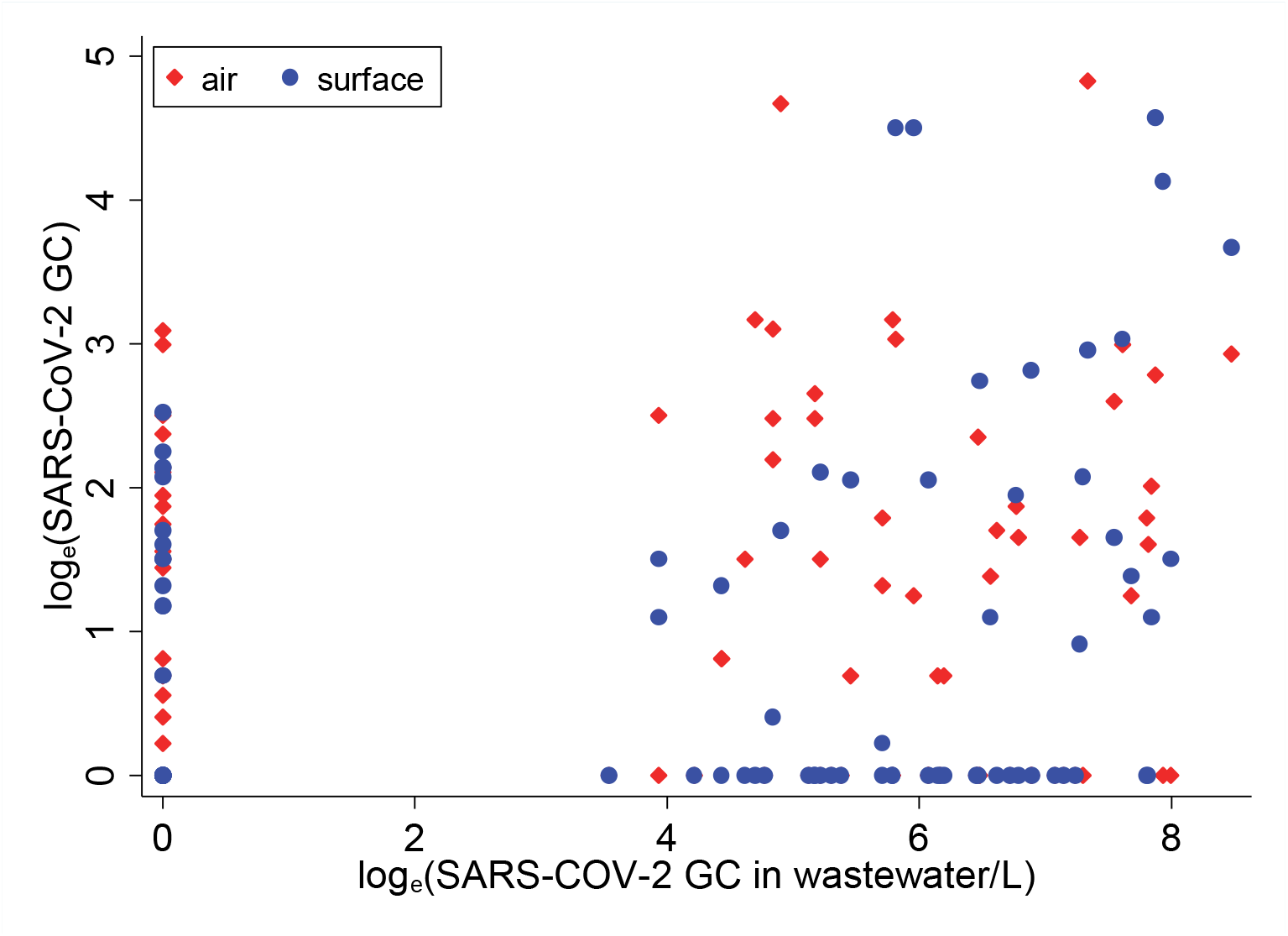
Distribution of one-day lagged moving average of SARS-CoV-2 in air and surface samples with respect to SARS-CoV-2 in wastewater samples, March-May 2021 (wastewater concentration on x-axis)

We conducted comprehensive environmental sampling of SARS-CoV-2, but human surveillance was limited to the residential dormitory selected for this study. On average, 2.3 students/day were tested on 44 of the 85 days during the study period. COVID-19 cases were detected on 11 of these 44 days of testing: 1 student tested positive during 9 of the days, and 2 students tested positive during the other 2 days. Given this gap in the COVID-19 surveillance, the comparison of COVID-19 cases and environmental SARS-CoV-2 was restricted to days of COVID-19 screening. We computed 1, 2 and 3 day lagged moving average of SARS-CoV-2 concentration in all collected samples. One-day lag moving averages of SARS-CoV-2 were above detection limit on 52 (63.4%), 40 (50.0%) and 57 (68.6%) of 83, 82, and 80 days in air, surface swab and wastewater samples, respectively. On the 11 days when COVID-19 cases were detected,

SARS-CoV-2 was detected in air, surface swab and wastewater samples on 6, 6, and 7 of these days respectively (Table S3). However, on 5, 5, and 4-days, air, surface swab, and wastewater samples were negative for SARS-CoV-2 on the same days when COVID-19 cases were detected in the building. The efficacy of SARS-CoV-2 detection in the 2-day lagged moving average of air, surface swab and wastewater samples to predict COVID-19 case(s) was 100%, 82.8% and 72.7%, respectively (Table 3). On 28 days when SARS-CoV-2 was detected in the three-day moving averages of air and wastewater samples, COVID-19 cases were not detected among the randomly selected residents screened for COVID-19.

When SARS-CoV-2 was detected in any of the three air, surface swab and wastewater samples on a given day, the efficacy of SARS-CoV-2 detection in 0, 1, 2, 3 and 4 day-lagged environmental samples to predict COVID-19 case(s) detection was 90.9%, 90.9%, 100%, 90.9% and 100% respectively. However, when the 1-day lagged moving average of any of the three types of samples was used, COVID-19 case(s) prediction rate using the 0 to 4 day lagged environmental average of SARS-CoV-2 increased to 100%, suggesting if one sample type fails to detect SARS-CoV-2, it can be detected in another type of sample (Table S4).

## DISCUSSION

SARS-CoV-2 detection in air, surface swab and wastewater samples in a building with clinically confirmed COVID-19 cases suggests that the genomic footprints of the virus, shed by infected individuals, can be traced in the environment. Our analysis further suggests that SARS-CoV-2 detection in environmental samples (air, surface swab and wastewater) predicted all clinically diagnosed COVID-19 cases in the selected dormitory 1-2 before the case-diagnosis date during the study period. Some of these findings are consistent with the emerging literature, which suggests association between COVID-19 case prediction with the aid of SARS-CoV-2 in wastewater samples [10, 14]. Daily environmental samples used in this research provide a novel insight into variations in COVID-19 case prediction with respect to changes in time-lagged SARS-CoV-2 detection in air, surface swab and wastewater samples. The 3-day lagged SARS-CoV-2 in the wastewater samples showed the strongest association with COVID-19 case detection, which is consistent with previous research. Sharkey et al. 2021 describe that the 4-day lagged SARS-CoV-2 in wastewater had the strongest association with COVID-19 cases [13]. This current study, for the first-time, shows association between COVID-19 cases and time-lagged SARS-CoV-2 detection in three different types of samples collected daily. In fact, the strongest association between COVID-19 diagnosis and SARS-CoV-2 in air and surface swabs was observed two days before COVID-19 case reporting. Thus, SARS-CoV-2 in these environmental samples can provide early warnings of an outbreak even at the scale of a building. A unique finding of this research is that if one environmental sample type was falsely negative (for SARS-CoV-2) it was detected in another sample type, suggesting that sampling air, surface swab and wastewater on the same day can dramatically improve the efficacy of environmental surveillance of infectious disease.

Our research has public health and policy implications. Consistent monitoring of air, swabbing high touch surfaces and wastewater analysis can detect genomic footprints of SARS-CoV-2. Thus, the detection of the virus in environmental samples several days prior to clinical diagnosis can guide timely interventions to reduce pathogen transmission. Unlike sentinel human screening, environmental monitoring is non-invasive and less costly [22, 23]. Moreover, asymptomatic individuals do not necessarily seek care and hence can be missed by clinical screening [24]. Environmental samples can help identify location- and time-specific cluster(s) of both symptomatic and asymptomatic cases several days prior to their clinical diagnosis. For example, strategic (active) air, surface and wastewater samplings in places such as airports, schools and malls can provide insight to the spread and potential outbreak of the disease at multiple scales, and trace sources of disease transmission.

Results of this research must be interpreted with caution due to the following limitations. First, some of the samples analyzed could be false negatives due to low concentrations of virus quantified from the sample or due to inhibition during sample preparation and analysis. Second, SARS-CoV-2 recovery from the samples can be subject to bias due to a low recovery rate from air, surface swab and wastewater samples. Third, COVID-19 case data can also be subject to bias because of the limited testing of students. For example, many days when SARS-CoV-2 was detected in the environmental samples, but COVID-19 cases were not reported. Routine daily testing of students was not implemented, especially during weekends. Finally, SARS-CoV-2 concentrations in the environmental samples were not adjusted for potential confounders, such as local meteorological conditions, ventilation and airborne particulate matter which have been shown to impact SARS-CoV-2 concentrations in air and on surface swabs [23, 25].

Despite these limitations this research shed light on the relevance of proactive environmental microbiome surveillance for emerging disease-causing pathogens and their management.

## Supporting information

Supplement Material and Methods and Detailed Results

## Data Availability

All environmental data can be made available upon a reasonable request to the corresponding author. However, clinical data are confidential and cannot be shared with any one outside our team.

## Author contributions

Conceptualization: NK, HS

Methodology: NK, HS, MS

Investigation: NK, HS, SK, SA, JC, MS, AM, WL, JTJ, EK, NS, RK, BS, SW

Visualization: NK

Funding acquisition: HS, SS, CM

Project administration: GG, HS, NK, SA, SK, DV

Supervision: NK, HS

Writing – original draft: NK, SK, SA

Writing – review & editing: HS, NK, TB, SA

## Data and materials availability

All masked environmental sample data can be made available upon a reasonable requested to the corresponding author. Human subject data on COVID-19 will not be available due to confidentiality reasons.

