## Supplement Material and Methods and Detailed Results for "COVID-19 Prediction using Genomic Footprint of SARS-CoV-2 in Air, Surface Swab and Wastewater Samples"

### METHODS AND MATERIALS

#### Experimental strategy

We used a prospective control design for this research. We selected a campus site (Y-leg of Lakeside Village [student dormitory], YLV) with corresponding wastewater and high traffic access points. YLV houses about 500 students and is serviced by two interconnected lobbies, C and D. This dorm is secure and can be accessed by students by swiping their university issued identification cards. Based on the swipe registry data, more than 5,000 individuals (mostly students) entered YLV in a seven-day period. The wastewater from the YLV drains to a designated manhole (K). Since there are two access points to YLV, we conducted daily (active) air sampling and surface swab sampling in both lobbies. Thus, 24h air samples and swab samples of high touch surfaces, namely elevator buttons, door handles and bars, corresponded to daily wastewater samples from manhole K (Figure S1). Random screening of students residing in the dormitory also occurred 2-3 times/week during the study period: March 2, 2021, to May 28, 2021. Methods of sample collection and analyses of these samples are detailed below. Daily air, surface swab and wastewater samples were collected between 8:00 AM and 9:00 AM Monday through Friday and 10:00 AM to 11:00 AM during weekends.

#### Sample Collection

**Wastewater** samples were collected from one manhole located at the Lakeside Village of the University of Miami corresponding to the undergraduate student dorm selected for this study (Figure S1). Daily sampling occurred from 2$^{nd}$ March 2021 to 28$^{th}$ May 2021. The samples were collected Mondays through Fridays at 8:30 AM ± 30 minutes, and Saturdays and Sundays at 10:30 AM ± 30 minutes.

For the first two weeks, only grab samples were collected using the ‘**bottle on chain’ approach**. In this method, a plastic wide-mouth bottle (HDPE), attached to a chain, was lowered in the manhole and the wastewater was collected. Once the sample was collected, it was transferred to another, smaller, sterile (HDPE) bottle to be transported to the laboratory. After two weeks, an **autosampler** (ISCO 6712) was installed and **composite samples** were collected along with the **grab samples**. Composite samples consisted of a mixture of hourly samples drawn in a 5-liter sterile bottle during a 24-hour cycle. The program for the autosampler was set one day prior to the collection of the composite sample. The volume of the sample collected hourly was adjusted between 175-225 mL, according to the flow of the wastewater through the manhole. When the wastewater flow was low or wastewater was not present in the manhole, the autosampler did not collect the sample. The autosampler recoded data on the number of samples collected, time of collection and volume sample for each collection.

Before the sample collection on a given day, the sampling log from the autosampler was retrieved. The log included the hour which the autosampler collected the sample, the number of samples collected, and their respective volumes. The 5-liter bottle with the composite sample was retrieved from the autosampler and replaced with a new sterile bottle. The total volume in the 5-liter bottle was noted. The wastewater collected in this bottle was mixed thoroughly and dispensed in a 500 mL plastic cup placed in a plastic container to avoid spillage. A stainless strainer with 3 mm circular openings was used to capture large solid particles in the wastewater. The sensor end of a pre-calibrated sonde (Xylem YSI Pro DSS) was submerged in the sample to measure sample characteristics, such as temperature, pH, salinity, dissolve oxygen, and turbidity. This sample was then transferred to a sterile 250 mL plastic bottle. The bottle was closed, spayed with isopropyl alcohol, dried, labelled with the respective sample ID (WGOKYYMMDD) with a permanent marker and placed in a zip lock bag. This sample was stored in an ice cooler and transported to our laboratory for processing on the same day. All instruments were rinsed with water, dried, sprayed with 70% isopropyl alcohol and again dried with a lint-free Kim wipe.

**Air pollution monitoring pumps with flowmeters (LMP AIR)** were installed in the Lobby C and Lobby D of the dorm selected for air sampling (Figure S2). The sampling set-up was customized so that the air was drawn to the air filter from both elevators. 1.42/$m^3$ air was drawn by the blower from two inlets. Using an L-connection, the air was branched (from the main stream of air flow) to the impactor, which was connected to a flow meter, and a vacuum pump drew air through the flowmeter at a set flow rate. Each day, a new sterile polycarbonate membrane filter **(Isopore, 0.4µm PC membrane, 47 mm**) with a 47 mm supporting cellulose pad was deployed in the impactor.

The pump was turned on and initial flow was set between 11 L ± 1 L. Starting date, time and initial flow rate were noted. The sample was collected next day, generally after 24 h ± 2h. We also deployed a real-time air pollution sensor that monitored airborne particle matter (PM) of three different sizes ≤ 1 µm, ≤ 2.5 µm and ≤ 10 µm ($PM_1$, $PM_{2.5}$ and $PM_{10}$, respectively), three gases CO, $NO_2$ and NH3, temperature and relative humidity. These data were also noted at the beginning and end of sampling period each day. The sensor also recorded these data every 3 minutes. After 24h, the flow was noted, pump was turned off, and the filter was retrieved from the impactor. The filter was rolled (cylindrically) and transferred to a 5 mL DNA/RNAase free conical tube. The impactor was cleaned with 70% isopropyl alcohol, wiped with a lint free Kim wipe, and new set of PC membrane filter and a 47 mm supporting cellulose pad was deployed for the next 24h. The retrieved samples from both Lobby C and Lobby D of the dorms were stored in an ice cooler, which were then transported to our laboratory for processing.

**Surface swab samples** were collected from high touch surfaces including elevators buttons, door handles and bars using sterile Polyester Swabs with Polystyrene Handle. Following the collection, the swabbed surfaces were wiped with alcohol wipes for sterilization. The swab tips were stored in 1.5 mL conical tubes in a cooler with ice packs and transported to our laboratory for processing.

**Disinfection practices** were prioritized throughout the process with oversight from the **University's Health and Safety Office**. **Safety protocol** was followed in the field, and all study team members used proper PPE, such as using disposable lab coats, disposable gloves, and reusable goggles. Masks and face shields were worn as part of COVID-19 social distancing protocols.

### Sample Processing

#### Bacteria by Culture

All wastewater samples were analyzed for the concentration of fecal indicator bacteria (FIB) to determine the concentration of human fecal input needed to standardize the concentration of SARS-CoV-2 with respect to estimated human bodily excretion. The wastewater sample was mixed, and an aliquot of 10 mL was removed and added in a sterile 15 mL centrifuge tube. 0.5 mL of the 10 mL (untreated) wastewater sample was added to 50 mL sterile phosphate buffered saline (PBS) to dilute the sample (100:1 dilution). After mixing the wastewater sample in the 50 mL PBS, 0.1 mL and 1 mL aliquots of the diluted samples were added in the 20 mL PBS in a sterile funnel and filtered through 47 mm grided membrane filters. Membrane filters were placed on the mFC agar plate and incubated for 24h ± 2h at 45°C **(Method 9222, APHA 2005)**. Agar plates were images and FIB colonies were counted using ImageJ [26].

**RNA Concentration - Electronegative Filtration**

The remaining 250 mL wastewater sample was used for the RNA concentration of SARS-CoV-2 using *electronegative filtration* described by **Method 9510, APHA 2017**. A magnetic stir bar was added to the sample to assist with maintaining homogeneity once placed on a magnetic plate. Heat inactivated (at 56°C for 15 minutes) 35 µL of Beta coronavirus OC43 with known number of copies, was added to 250 mL of wastewater sample, which served as a positive control for RNA extraction and its quantitation. To assist with adhesion to the filter, 2.35 mL of magnesium chloride was added per sample then drop by drop 10% HCl (acid) was included to acidify the sample to a target pH value between 4.5-3.5. The number of HCl droplets added in the sample and the final pH value of the homogenized sample were noted. 40 mL ± 10 mL (depending on the presence of solid material dissolved in the water) was filtered through an electronegative membrane filter (0.45µm pore size, 47mm diameter). The filter was rolled conically and placed in an RNase/DNase free 5 mL conical tube. 1 mL of the DNA/RNA shield (ZYMO) was added to the 5 mL tube. The tube was vortexed to rinse of the concentrated wastewater water sample, and then centrifuged. The concentrated was mixed by pipetting and a 250 µL aliquot was used for subsequent RNA extraction.

**RNA Concentration - Air Filter Samples**

Heat inactivated (56°C for 15 minutes) 35 µLof Beta coronavirus OC43 with a known concentration between $10^5$ and $10^6$ genomic copies (gc)/L was added as droplets to the filters while still in the 5 mL the conical tube. 500 µL of DNA/RNA shield was then added around the top of the filters to collect any particles on the filter. This step was repeated, and air filter was submerged in 1 mL of DNA/RNA shield. The tube was vortexed and centrifuged, and the concentrate was mixed by pipetting and a 250 µL aliquot of the concentrated was used for RNA extraction.

**RNA Concentration - Surface Swab Samples**

Heat inactivated (56°C for 15 minutes) 35 µL of Beta coronavirus OC43 with a known concentration between $10^5$ and $10^6$ gc/L was added as droplets to the swabs while still in the 1.5ml conical tube. 1 mL of DNA/RNA shield was then added around in the tube. The tube was vortexed and centrifuged. The concentrate was mixed by pipetting and a 250 µL aliquot of the concentrate was used to for RNA extraction.

**SARS-CoV-2 RNA Extraction Process**

The nucleic acid extraction process consists of three processes: isolation, purification, and concentration (Figure S3). We used a *QuickRNA*-Viral 96 Kit from Zymo Research Inc. for RNA extraction from all three sample types following their R1040/R1041 protocol. We added 500 µL of Viral RNA binding buffer to the concentrates of 250 µL wastewater, air filter, and surface swab samples previously prepared in 1 mL DNA/RNA shield. This reagent facilitates particle lysis and binding of RNA from other biological liquids, such as urine, in the wastewater. The samples were then centrifuged and transferred to columns and collections tubes. Next, 500 µL of wash buffer was added to the column before centrifuging. This was repeated twice. This "wash" removes proteins, salts, and other contaminants from the sample. 500 µL of 100% ethanol was then added to the samples before centrifuging, which allowed the RNA to precipitate since nucleic acids are insoluble in ethanol. Finally, we added 15 µL of RNase-free water to the columns. After waiting for a minute, and the columns were centrifuged and RNA from the columns were collected in the 1.5 mL RNase free conical tubes for the follow up qPCR analysis.

**RNA Quantification**

The qPCR method utilized here was Volcano Second Generation (V2G) as described in Sharkey et al. [13]; aliquots of purified RNA were used in singleplex reactions to quantitate the SARS-CoV-2 N3 nucleocapsid gene. An advantage of the V2G-qPCR method over the more mainstream RT-qPCR is that it can read both RNA and DNA templates which eliminates the prior cDNA synthesis step [20]. According to research, the coronavirus nucleocapsid protein assists in the replication and transcription of viral RNA and interferes with cell-cycle processes of host cells, and as a result, plays a critical role in SARS-CoV-2 pathogenesis. The nucleocapsid proteins of many coronaviruses are immunogenic and expressed abundantly during infection [27]. Detection, quantification, and analysis of its presence in wastewater, air filter, and surface swab samples have allowed for relatively accurate prediction of SARS-CoV-2 cases in a student dormitory and has correlated with reported cases at the University of Miami.

**qPCR Process**

20 μL of HIV-1 RNA spike is added to all samples before the qPCR process to assess PCR inhibition. A master mix protocol, created in-house based on the number of samples being run, and involved the following volumes and reagents for SARS-CoV-2 detection:

| qPCR Master Mix Reagents | Volume per Reaction |
|---|---|
| RNase-free water | 17.7 μL |
| 5X Volcano (2G) Buffer | 6.6 μL |
| 10 mM dNTPs | 0.6 μL |
| 5 units/μL anti-Taq antibody | 0.15 μL |
| 5 units/μL Volcano (2G) Polymerase | 0.3 μL |
| 20 μM CV3b/f primer | 0.75 μL |
| 20 μM CV3c/r primer | 0.75 μL |
| 100 μM CV3 probe | 0.075 μL |
| 400X Rox | 0.075 μL |

Utilizing a 96-well plate (BioRad Hard Shell #HSP9601), 27 μL of master mix was added to individual wells before the RNA of samples, or any controls were inserted. 3 μL was utilized for all inputs to the plate which included sample RNA for "unknown" wells, nuclease-free water for no template, or negative, controls (NTCs), and Twist positive standards. To utilize a standard curve, 3 μL of SARS-NC product ranging from $10^1$ to $10^5$ cp/uL were also added. The plate was sealed by firmly pressing on an adhesive Microseal B (BioRad #MSB1001), and briefly centrifuged prior to being loaded in the machine. A BioRad CFX-Connect instrument was used for real time results of SARS-CoV-2 detection.

This analysis was repeated for the viral recovery control OC43 with a known concentration between $10^5$ and $10^6$ gc/L following its addition to all sample types (air, swab, wastewater). Utilizing a recovery control allowed for a downstream percent recovery to be calculated per sample as a surrogate for estimating the viral degradation of SARS-CoV-2. For OC43 detection by qPCR, a similar master mix was created in-house based on the number of samples being run, except for the target-specific primers and probe in which 20 μM OC43 f/r primer's and a 100 μM OC43 probe were utilized prior to analysis with qPCR.

Similarly to before, 27 μL of the master mix was added to individual wells of a 96-well plate (BioRad Hard Shell #HSP9601) before 3 ul of sample was included to the wells indicating “unknowns”, a 3 μL volume was also utilized of nuclease-free water for NTCs as well as for positive Twist control standards. The standards included for OC43 analysis ranging from $10^1$ to $10^5$ cp/uL consisted of pre-quantified PCR-amplified OC43 product. The plate was sealed by firmly pressing on an adhesive Microseal B (BioRad #MSB1001), and briefly centrifuged prior to its placement in the machine. A BioRad CFX-Connect instrument was also used for real time results of OC43 detection.

**COVID-19 surveillance** of the students residing at the YLV dormitory consisted of random screenings 2-3 days/week using nasal swabs which were analyzed by RT-PCR. These anonymous data on the total number of tests (performed) and COVID-19 cases by date were acquired from the UM administration.

**Statistical analysis** conducted were descriptive analyses of aggregated and disaggregated data. Data of air and surface swab samples from both lobbies were aggregated to compare them with the data from the wastewater samples. $\chi^2$ tests were performed to assess statistical differences across groups and a p-value of 0.05 or below was considered as significant.

### FIGURES AND TABLES

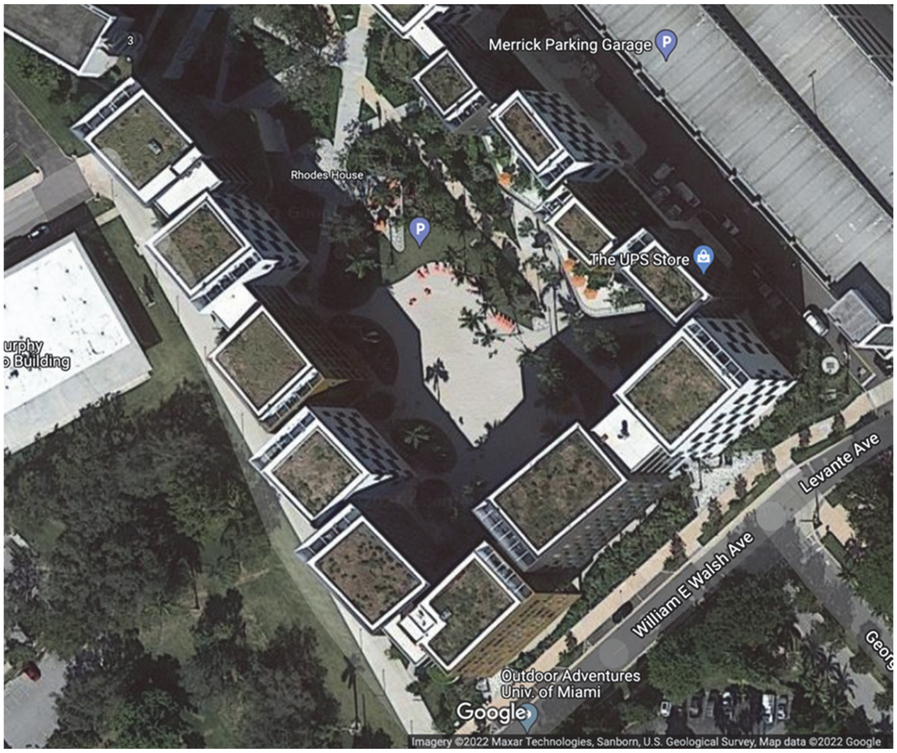


**Figure S1:** Map showing the Y-leg of the Lakeside Village indicating wastewater sample collection site, Lobby C, and Lobby D where air filter, and surface swab samples were collected.

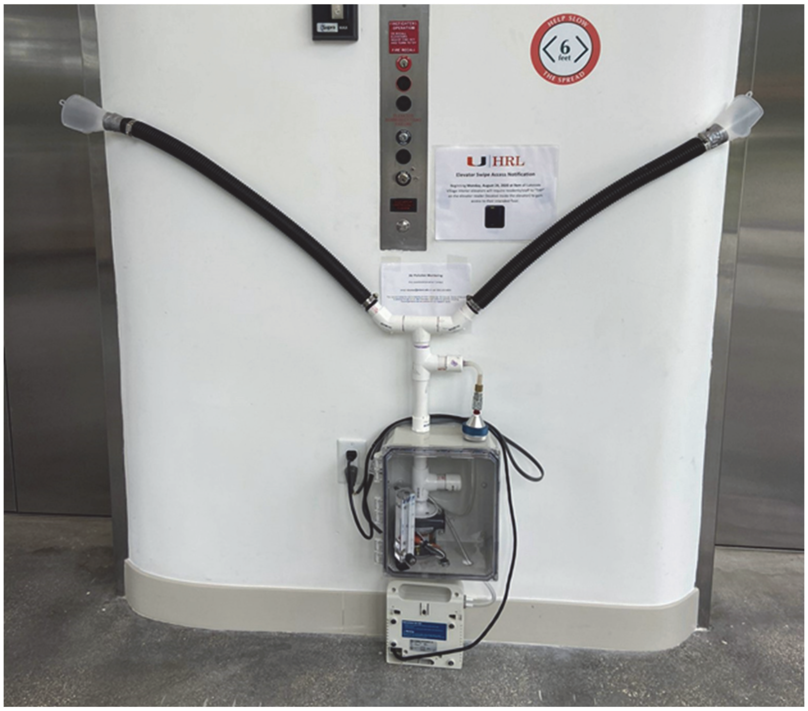


**Figure S2:** Active air sampling setup in lobby of Lakeside Village. Sterile 47mm polycarbonate membrane filters with a pore size of 0.4 μm were used in our customized impactors, connected with a vacuum pump that ran in 25h increments at a flow rate of 10-12 l/minute to collect particles and aerosols within the lobby.

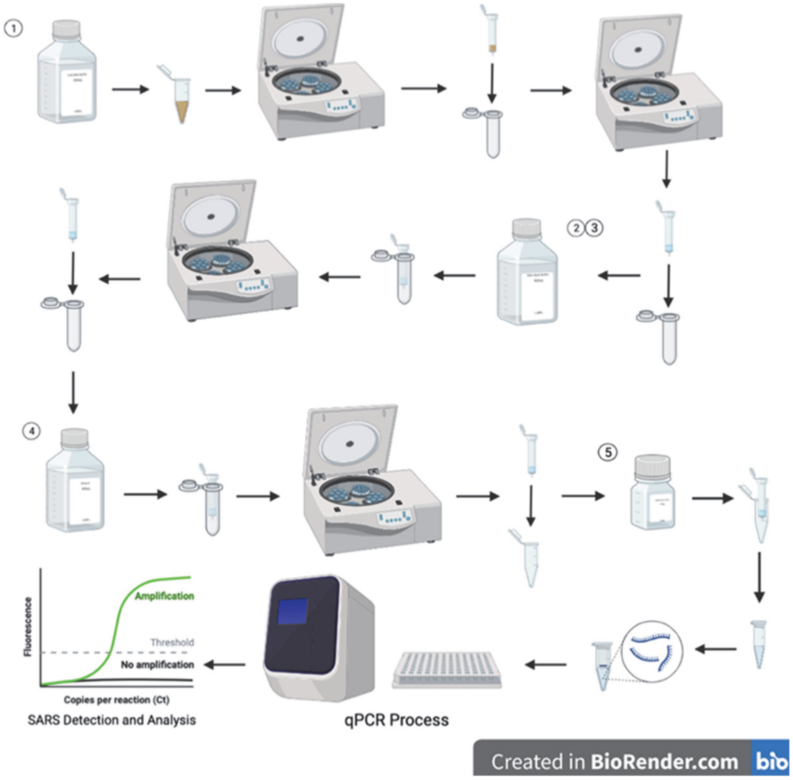


**Figure S3***:* SARS-CoV-2 extraction process showing the five steps to prepare for qPCR process.

| Ref | Year | Findings | Detailed Findings | # samples | # positive cases | Positivity rate % | Media | Study Site |
|---|---|---|---|---|---|---|---|---|
| [28] | 2020 | SARS-CoV-2 decays faster when temperature/humidity are increased | "At room temperature (24°C), virus half-life ranged from 6.3 to 18.6 h depending on the relative humidity but was reduced to 1.0 to 8.9 h when the temperature was increased to 35°C …findings suggest that a potential for fomite transmission may persist for hours to days in indoor environments…" | | N/A | N/A | Virus containing droplets (stainless steel and plastic) | Fort Detrick, Maryland (NBACC) |
| [29] | 2020 | SARS-CoV-2 more stable on plastic and stainless steel (detected 72 hours after application) | "…results indicate that aerosol and fomite transmission of SARS-CoV-2 is plausible, since the virus can remain viable and infectious in aerosols for hours and on surfaces up to days…" | | | | Surface | |
| [30] | 2020 | Small particles containing SARS-CoV-2 can diffuse indoors even up to 10m from source | "…The available information on the SARS-COV-2 spreading supports the hypothesis of airborne diffusion of infected droplets from person to person at a distance greater than two meters (6 feet). The inter-personal distance of 2 m can | N/A | N/A | N/A | Air | Lit review |

| | | | | | | | | |
|---|---|---|---|---|---|---|---|---|
| | | | be reasonably considered as an effective protection only if everybody wears face masks in daily life activities.” | | | | | |
| [10] | 2021 | WBE as early warning | “…accurate diagnostic for new cases…with 82.0% positive predictive value…” | 111 | 91 | 82% | Wastewater | University of Arizona (dorms) |
| [11] | 2021 | Wastewater as emerging tool to help identify covid outbreaks | “…analysis suggests that wastewater monitoring at colleges requires consideration of information needs, local sewage infrastructure, resources for sampling and analysis, college and community dynamics, approaches to interpretation and communication of results, and follow-up actions.” | Varied by school | | | Wastewater | Compiled from 25 universities |
| [12] | 2021 | Wastewater was good predictor | “…temporal trends of SARS-CoV-2 in wastewater samples mirrored trends in COVID-19 cases detected on campus… N1 and N2 genes were significant predictors of COVID-19 cases in dormitories, and the N2 gene was significantly correlated with the number of detected COVID-19 cases in dormitories.” | 117 | | | Wastewater | Tulane University |
| [14] | 2021 | Wastewater better alternative to pooled/random testing and was able to identify cases; likely applicable to future pandemics in group settings | “…wastewater surveillance enabled the identification of asymptomatic COVID-19 cases that were not detected by other [testing]…able to detect single symptomatic individuals in dorms with resident populations of 150-200…” | 332 | | By sample | Wastewater | University of North Carolina at Charlotte (dorms) |
| [15] | 2020 | High touch surface contamination and air contamination after first week of illness | “CoV-2 PCR-positive particles of sizes >4 microm and 1-4 microm in two rooms, despite these rooms having 12 air changes per hour.” | 15 patients | 10 patients | 67% showed high touch contamination | Surface and air samples | Singapore (NCIF ICU) |
| [22] | 2021 | Wastewater as identifier | “SARS-CoV-2 RNA fragments have been isolated from numerous wastewater treatment works, septic tanks, sewers, hospital wastewater treatment systems, and environmental discharge points[7] and reported to predate the clinical diagnosis of cases,[9,10] raising the potential for its use...” | N/A | N/A | N/A | Wastewater | Lit review |
| [23] | 2021 | Noninvasive environmental surveillance useful as long-term method for monitoring of covid and future diseases | “SARS-CoV-2 can be detected in bulk floor dust collected from rooms housing infected individuals. This analysis suggests that dust may be a useful and efficient matrix for routine surveillance of viral disease.” | ~59.6 | 67% | 89% bulk dust samples positive; 55% of surface swab samples positive | Surface swab, passive sampler, and bulk floor dust | Ohio State University (2 houses; isolation dorms) |

| | | | | | | | | |
|---|---|---|---|---|---|---|---|---|
| [24] | 2021 | Surface, aerosol, and wastewater all reliable methods of Covid surveillance; sludge another option; nucleic acid analysis, isothermal amplification, CRISPR, mass spectrometry, etc. other options for detection methods | "The characteristics of anti-interference and reliable quantification make the ddPCR technique a good choice for detecting CoVs from various environmental matrices… to address current challenges in detection techniques, further researches towards multiple chemistry-related disciplines have to be strengthened…(1) the guarantee of sensitivity and reliability… (2) Quantitative analysis of CoVs… (3)The confirmation of viability…" | N/A | N/A | N/A | Environmenta l | Lit review |
| [25] | 2020 | PM (particulate matter) plays role in providing surfaces for pathogen transmission; chemical composition and mass of PM is highly variable and dependent on things like geography, season, emissions, etc. | "Virus deposition rates positively correlated with organic aerosols of size <0.7 μm, implying longer residence time and further dispersion of viruses. Hence PM can play a vital part in virus transmission and survival. " | N/A | N/A | N/A | Water and air | Lit review |
| [31] | 2021 | Virus can accumulate and be inhaled more easily in indoor spaces due to poor ventilation; CO2 monitors are possible solution; experiments unethical because it would be exposing people to virus | "…unclear how much ventilation is needed to reduce infection rates to an acceptable level… The precise infection dose for SARS-CoV-2 is unknown." | | | | Air | Lit review |
| [32] | 2021 | Viral RNA in feces | "The need for the development of robust sampling strategies and subsequent detection methodologies and challenges for developing countries.." | N/A | N/A | N/A | Wastewater | Lit review |
| [33] | 2020 | Covid found in patient's rooms at hospitals but doesn't indicate whether it is airborne or transmitted by droplets | "…most of the used samplers such as PTFE filters, gelatin filers and cyclones showed suitable performance for trapping SARS-Co and MERS-Cov viruses followed by PCR analysis." | | | | Air | |
| [34] | 2021 | Crowd sourcing can be a useful methodology to collect microbiome samples from public places. | Crowd sourcing a useful methodology for collecting environmental samples from public places. Samples were sensitive and resistant to inhibitors present environmental samples. | 4,080 | | | Surface swabs | San Diego, California |
| [35] | 2021 | Wastewater was good predictor | "SARS-CoV-2 N gene daily loads strongly correlated with the number of cases | 185 | | | Wastewater | Catalonia, Spain |

| | | | | | | | | |
|---|---|---|---|---|---|---|---|---|
| | | | diagnosed one week after sampling.” | | | | | |
| [36] | 2021 | Association between Covid cases and particulate matter levels; this has implications for policymakers, clinicians, public health authorities | “They found an 8% increase in the Covid-19 mortality rate (95% CI 2%, 15%) per unit increase of $PM_{2.5}$.” | | | | Air (outdoors) | Multiple locations globally (outdoors) |
| [37] | 2020 | Increasing evidence for droplet-aerosol transmission as a route for infection | “…$PM_{2.5}$ may provide a good platform to “shade” and “carry” the SARS-CoV-2 during atmospheric transport. Thus, $PM_{2.5}$ containing SARS-CoV-2 could be a direct transmission model in a highly polluted area.” | N/A | N/A | N/A | Air (outdoors) | Lit review |
| [38] | 2020 | Asymptomatic carriers can still contaminate surroundings; cleaning and negative pressure rooms ideal for isolation | “…in a single room occupied by an asymptomatic patient, four sites were SARS-CoV-2 positive, highlighting that asymptomatic COVID-19 patients do contaminate their surroundings.” | 112 | 44 | 39.3 | Surface swab | Chengdu, China (non-ICU isolation ward) |

**Table S1:** Literature on environmental SARS-CoV-2 and COVID-19 surveillance

| | ***SARS-CoV-2 Detection*** | **Surface swab sample detection** | | |
|---|---|---|---|---|
| | | NO | YES | **TOTAL** |
| **Wastewater sample detection (NO)** | | | | |
| **Air sample detection** | NO | 5 (20.8) | 2 (8.3) | **7 (29.2)** |
| | YES | 5 (20.8) | 12 (50.0) | **17 (70.8)** |
| | TOTAL | 10 (41.7) | 14 (58.3) | **24 (100)** |
| **Wastewater sample detection (YES)** | | | | |
| **Air sample detection** | NO | 14 (25.9) | 7 (13.0) | **21 (38.9)** |
| | YES | 14 (25.9) | 19 (35.2) | **33 (61.1)** |
| | TOTAL | 28 (51.9) | 26 (48.1) | **54 (100)** |

**Table S3:** Number of days SARS-CoV-2 detection in air, surface swab and wastewater samples, March-May 2021 (% days in parenthesis).

| COVID-19 detection | Same day | | 1-day lag | | 2-day lag | | 3-day lag | |
|---|---|---|---|---|---|---|---|---|
| | No | Yes | No | Yes | No | Yes | No | Yes |
| **Air Samples** | | | | | | | | |
| No | 15 (75.0) | 17 (73.9) | 12 (92.3) | 20 (66.7) | 8 (100) | 25 (69.4) | 5 (100) | 28 (71.8) |
| Yes | 5 (25.0) | 6 (26.1) | 1 (7.7) | 10 (33.3) | 0 (0.0) | 11 (30.6) | 0 (0.0) | 11 (28.2) |
| COVID-19 prediction rate (%)* | 54 | | 90.9 | | 100 | | 100 | |
| % overall agreement** | 48.8 | | 51.2 | | 43.2 | | 36.4 | |
| **Surface Swab Samples** | | | | | | | | |
| No | 18 (78.3) | 11 (64.7) | 12 (80.0) | 17 (68.0) | 12 (85.7) | 21 (70.0) | 12 (85.7) | 21 (70.0) |
| Yes | 5 (21.7) | 6 (35.3) | 3 (20.0) | 8 (32.0) | 2 (14.3) | 9 (30.0) | 2 (14.3) | 9 (30.0) |
| COVID-19 prediction rate (%)* | 54 | | 72.7 | | 81.8 | | 81.8 | |
| % overall agreement** | 60.0 | | 50.0 | | 47.7 | | 47.7 | |
| **Wastewater Samples** | | | | | | | | |
| No | 19 (82.6) | 12 (63.2) | 10 (71.4) | 21 (75.0) | 8 (72.7) | 25 (75.8) | 5 (83.3) | 28 (73.7) |
| Yes | 4 (17.4) | 7 (36.8) | 4 (28.6) | 7 (25.0) | 3 (27.3) | 8 (24.2) | 1 (16.7) | 10 (26.3) |
| COVID-19 prediction rate (%)* | 63.6 | | 63.6 | | 72.7 | | 90.9 | |
| % overall agreement** | 61.9 | | 40.5 | | 36.4 | | 34.1 | |

* = 100 x ((# day when COVID-19 case(s) were detected / # days SARS-CoV-2 detection in the environmental samples when COVID-19 case(s) were detected)

** refers to agreement in frequency of days when COVID-19 and time-lagged environmental SARS-CoV-2 detection status matched, i.e. 100 x (# days when COVID19 and SARS-CoV-2 were negative + # days when both were positive/ total number of days of testing)

**Table S3:** Number of days COVID-19 case reporting with respect to time-lagged SARS-CoV-2 detection in environmental samples (column % in parenthesis).

| Date | Number of COVID-19 cases | Air SARS-CoV-2 GC / m$^3$ | | | Surface SARS-CoV-2 GC / m$^2$ | | | Wastewater SARS-CoV-2 GC / L | | |
|---|---|---|---|---|---|---|---|---|---|---|
| | | Lag-1 | Lag-2 | Lag-3 | Lag-1 | Lag-2 | Lag-3 | Lag-1 | Lag-2 | Lag-3 |
| 3/9/2021 | 1 | 4.0 | 3.3 | 2.5 | ND | ND | ND | 2,475 | 1,700 | 1,481 |
| 3/15/2021 | 1 | 5.5 | 7.7 | 9.4 | ND | ND | ND | ND | ND | 88 |
| 3/27/2021 | 1 | 17.8 | 11.8 | 8.9 | 38.3 | 27.5 | 23.4 | 4,775 | 3,183 | 2,388 |
| 4/6/2021 | 1 | 7.5 | 15.5 | 14.4 | 7.0 | 4.7 | 3.8 | ND | ND | ND |
| 4/8/2021 | 1 | 0.5 | 3.2 | 4.0 | 2.8 | 4.2 | 4.9 | ND | ND | ND |
| 4/9/2021 | 1 | 1.3 | 1.2 | 3.0 | 2.8 | 1.8 | 3.1 | 83 | 56 | 42 |
| 4/13/2021 | 1 | 19.8 | 15.7 | 11.8 | 89.5 | 61.2 | 45.9 | 333 | 222 | 167 |
| 4/19/2021 | 2 | 3.5 | 8.3 | 8.0 | 7.3 | 7.7 | 5.8 | 183 | 122 | 1,033 |
| 4/20/2021 | 1 | 2.8 | 2.3 | 6.3 | ND | 4.8 | 5.8 | 300 | 200 | 150 |
| 4/23/2021 | 2 | 1.0 | 0.7 | 0.5 | 4.5 | 3.0 | 2.3 | ND | 844 | 692 |
| 4/27/2021 | 1 | ND | 12.7 | 62.0 | 15.8 | 22.7 | 17.0 | 975 | 1,583 | 1,254 |
| ------------------------------------------------------------------------------- |  |  |  |  |  |  |  |  |  |  |
| # days COVID-19+ | | 90.9 | 100 | 100 | 72.7 | 81.8 | 81.8 | 63.6 | 72.7 | 72.7 |
| # days COVID-19- | | 63.6 | 72.7 | 84.8 | 51.5 | 51.5 | 51.5 | 66.7 | 72.7 | 84.8 |

**Table S4:** COVID-19 cases and time-lagged SARS-CoV-2 GC in environmental samples.
